# Anti-HBsAg antibody mAb19-LS enhances antiviral immunity in humans with chronic hepatitis B

**DOI:** 10.64898/2026.07.03.26357226

**Authors:** Zijun Wang, Mary Tenuta, Han Ngoc Le, Siri Nana Halling Svensgaard, Gabriela S. Silva Santos, Daniel B. Reeves, Gaëlle Breton, Vanessa Igbokwe, Andre Moraes Nicola, Katrina Millard, Sidsel Dahl Winther Andersen, Henriette Graversen, Caroline Zöllner, Martin Kluge, Rachel Scheck, Nina Weis, Isik S. Johansen, Deanna Dong, Brianna Hernandez, Irina Shimeliovich, Juan Dizon, Valeska Vieira, Frank Fabris, Magdalena Schwarzmüller, Pinkus Tober-Lau, David Hillus, Münevver Demir, Anna Gazumyan, Frank Tacke, Leif Erik Sander, Florian Kurth, Thomas A. Rasmussen, Hector Ye, Calvin Pan, Ira Jacobson, Qiao Wang, Jesper Damsgaard Gunst, Christian Gaebler, Ole S. Søgaard, Marina Caskey, Michel Nussenzweig

**Affiliations:** Laboratory of Molecular Immunology, The Rockefeller University, New York, NY, USA; Department of Dermatology, Laboratory of Precision Dermatology and Molecular Immunology of the Biomaterials and Regenerative Medicine Institute, Shanghai Ninth People’s Hospital, Shanghai Jiao Tong University School of Medicine, Shanghai, China; Laboratory of Translational Immunology of Viral Infections, Department of Infectious Diseases and Critical Care Medicine, Charité–Universitätsmedizin Berlin, Berlin, Germany; Berlin Institute of Health, Berlin, Germany; Department of Infectious Diseases, Aarhus University Hospital, Aarhus, Denmark; Vaccine and Infectious Disease Division, Fred Hutchinson Cancer Center, Seattle, WA, USA; Faculty of Medicine, University of Brasilia, Brasilia, DF, Brazil; Department of Hepatology and Gastroenterology, Campus Virchow-Klinikum (CVK) and Campus Charité Mitte (CCM), Charité–Universitätsmedizin Berlin, Berlin, Germany; Department of Infectious Diseases, Copenhagen University Hospital - Amager and Hvidovre, Hvidovre, Denmark; Department of Clinical Medicine, Faculty of Health and Medical Sciences, University of Copenhagen, Copenhagen, Denmark; Department of Infectious Diseases, Odense University Hospital, Odense, Denmark; Department of Infectious Diseases and Critical Care Medicine, Charité–Universitätsmedizin Berlin, Berlin, Germany; Department of Clinical Medicine, Aarhus University, Aarhus, Denmark; Medical Procare PLC, New York, NY USA; Department of Gastroenterology and Hepatology, New York University Langone Health, New York, NY USA; Department of Medicine, New York University Grossman School of Medicine, New York, NY USA; Key Laboratory of Medical Molecular Virology, Shanghai Institute of Infectious Disease and Biosecurity, School of Basic Medical Sciences, Fudan University, Shanghai 200032, China; Howard Hughes Medical Institute, The Rockefeller University, New York, NY, USA

## Abstract

Chronic infection with hepatitis B virus (HBV) is characterized by persistent expression of hepatitis B surface antigen (HBsAg), which is associated with profound immune tolerance. Although nucleos(t)ide analogue therapy effectively suppresses viral replication, it neither eliminates HBV nor reverses virus-specific immune dysfunction. Here, we report the results of two parallel first-in-human, dose-escalation studies evaluating a single infusion of mAb19-LS, a long-acting IgG1 monoclonal antibody targeting HBsAg, in individuals with chronic HBV infection receiving nucleos(t)ide analogue therapy. mAb19-LS was generally safe and well tolerated and induced a mean 11-fold increase in antigen clearance. The magnitude and duration of HBsAg suppression were dependent on both baseline antigen levels and mAb19-LS dose, with suppression maintained for more than 36 weeks in individuals receiving the highest dose. Reduction of circulating HBsAg was associated with uptake of HBsAg–IgG immune complexes by monocytes and dendritic cells and inflammatory reprogramming of these antigen-presenting cells. Notably, proliferation of both CD4^+^ and CD8^+^ T cells, as well as interferon-γ and TNF-α production in response to HBV antigens, were significantly increased 24 weeks after infusion. Together, these findings demonstrate that mAb19-LS is generally safe and effectively accelerates HBsAg clearance while activating antigen presenting cells and enhancing antiviral T cell responses.

## Introduction

Hepatitis B virus (HBV) infection remains a major global health burden and a leading cause of cirrhosis and hepatocellular carcinoma^1–3^. Whereas the majority of immunocompetent adults clear acute infection spontaneously, perinatal or early childhood exposure frequently results in chronic infection, underscoring the role of immune tolerance in disease persistence. Although nucleos(t)ide analogues effectively suppress viral replication, they do not eliminate hepatitis B surface antigen (HBsAg) production in most individuals^4^. Resolution of infection is mediated by a combination of T cell dependent noncytolytic interferon-γ (IFN-γ) and tumor necrosis factor (TNF-α) mediated mechanisms and direct cytolysis^5–8^.

During chronic infection, persistent exposure to high levels of HBsAg are associated with impaired dendritic cell (DC) function and dysfunctional HBV-specific T cells^9–12^. DCs exhibit impaired antigen presentation and increased inhibitory signaling that may limit effective priming of virus-specific T cells^9,11,12^. HBV-specific CD8⁺ and CD4⁺ T cells exhibit diminished proliferative capacity and reduced production of antiviral cytokines such as IFN-γ and TNF- α^10,13,14^. HBV-specific T cell dysfunction is heterogeneous and does not conform to current paradigms of terminal exhaustion but instead comprises several different potentially reversible differentiation states^10–12,15,16^. Finally, chronically infected individuals show low levels of anti-HBsAg IgG1 and relatively elevated levels of immunosuppressive IgG4^17–20^. Mechanistic studies in animal models further illustrate the complex relationship between antigen persistence and tolerance^7,10,12,21^.

Whether antibody-mediated immune pressure can impact dysfunctional DCs and HBV-specific T cells in humans remains unclear^22–24^. Here we report the results of two parallel first-in-human, dose-escalation phase I studies of anti-HBsAg mAb19-LS in patients with chronic HBV infection receiving nucleos(t)ide analogue antiviral therapy^25^.

## Results

To determine whether passively administered antibodies can impact anti-HBV immunity, we performed two parallel dose-escalation phase I clinical trials of mAb19-LS in individuals with chronic HBV infection receiving suppressive nucleos(t)ide analogue therapy (NCT05856890, EUCT number: 2023-508444-22-00 and NCT06668727). Participants received a single intravenous dose of 1, 3, 10 or 30 mg/kg mAb19-LS or placebo (n=16 and n=5, respectively; see Methods section). Here, we present data on these cohorts through 36 weeks after dosing (Fig. 1a). Of the 21 participants enrolled, 16 were male and 5 were female, with a mean age of 46 (range 20-70) years. Baseline demographic and clinical characteristics were comparable across cohorts (Supplementary Table 1, Supplementary Fig. 1). Most reported adverse events were of grade 1 severity, and no serious adverse events were reported (Supplementary Table 2,3). Transient infusion reactions with grade 1 and 2 systemic symptoms occurred in 3 of 15 participants who received mAb19-LS at 1, 3 and 30 mg/kg doses and had baseline HBsAg of approximately 4.0 log10 IU/mL or higher. Symptoms occurred during the infusion and were short lived, generally resolving within hours following treatment with non-steroidal anti-inflammatory and/or anti-histaminic drugs. Of those three participants, one was excluded from further analysis because the infusion was discontinued prior to completion. Another experienced grade 2 elevation in transaminases, which resolved within 2 weeks (Supplementary Fig. 2b). All three participants experienced grade 2 or 3 lymphopenia that resolved by day 1 (Supplementary Fig. 2a,b). Despite these events, dose-escalation proceeded according to protocol and following review by an external safety monitoring committee, reached the maximum dose of 30 mg/kg. mAb19-LS infusion induced a rapid initial reduction in circulating HBsAg reaching a mean nadir of 1.1-1.6 log10 IU/mL below baseline between weeks 1 and 4 for each of the 4 dosing groups (Fig. 1b-e). Thus, while the magnitude of the initial decline was similar across doses, the duration of suppression was dependent on mAb19-LS dose and baseline antigen levels, with sustained reduction after 36 weeks in the 30 mg/kg cohort (Supplementary Fig. 3,4). We developed mechanistic mathematical models to integrate longitudinal mAb19-LS concentrations and HBsAg levels. By fitting these models to the data from all participants using population nonlinear mixed effects modeling, we estimated the population mean HBsAg half-life at baseline was 1.6 days (95% CI: 0.9-10.6), which is in general agreement with prior estimates ^26^. Among 13 individuals who reached a new lower equilibrium after mAb-19-LS infusion, model estimated clearance rates were increased by a mean of 11-fold (95% CI range 4-21-fold), dropping their median baseline HBsAg half-life from 38.4 hours to a median half-life of 3.9 hours at equilibrium (Fig. 1f).

**Fig. 1:**
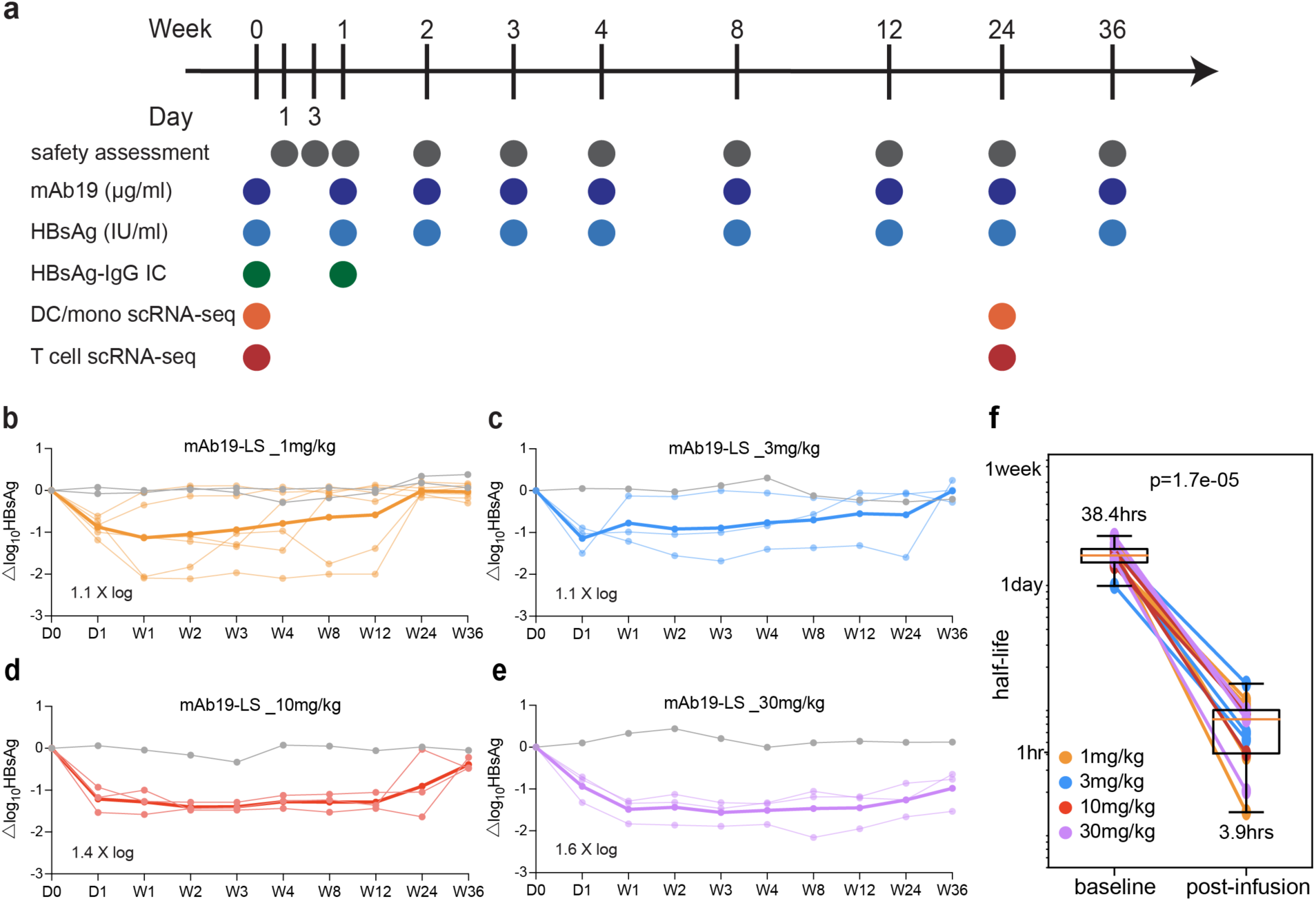
Reduction of circulating HBsAg following mAb19-LS infusion. **a**, Study design. Participants with chronic hepatitis B receiving suppressive nucleos(t)ide analogue therapy received a single intravenous infusion of 1 (n=6), 3 (n=3), 10 (n=3), or 30 (n=3) mg/kg mAb19-LS or placebo and were followed for 36 weeks. **b-e** Longitudinal HBsAg levels in individual participants across dose cohorts. Fine lines represent individual trajectories; Bold lines indicate median values; grey lines show individuals that received placebo. **f**, Estimated HBsAg half-life at baseline and at the post-treatment equilibrium state for individual mAb19-LS recipients that reached equilibrium before rebound (n=13, including PUB- 003, 004, 007, 008, 101,102, 105, 107, 112, 202, 203, 204 and 207). Each dot represents one participant, and paired points are connected by lines. Box plots indicate mean and interquartile range. The y-axis is shown on a logarithmic scale. The p-value was determined by two-sided paired comparison as indicated.

To determine whether decreased circulating HBsAg is associated with formation of circulating immune complexes, we quantified the proportion of HBsAg in serum bound to IgG using Protein G (Fig. 2a). At baseline, only a variable fraction of 0.5-64% (mean=18.5%) of the circulating HBsAg was IgG-associated, and this increased significantly to 67-100% (mean=90%) after mAb19-LS infusion (p<0.001, Fig. 2b). While immune complex levels declined after rebound, the fraction of IgG-associated HBsAg remained significantly elevated compared to baseline, possibly reflecting residual mAb19-LS (Fig. 2b). To examine the nature of the HBsAg associated antibodies, we also collected immune complexes from serum by polyethylene glycol precipitation and analyzed the associated antibody isotypes by ELISA (Fig. 2c-e, Supplementary Fig. 5). We found the expected increase in IgG1-containing HBsAg immune complexes and a small but significant decrease in IgG4 complexes, likely due to competition between mAb19-LS and IgG4 (Fig. 2d,e). Thus, mAb19-LS infusion traps nearly all circulating HBsAg in IgG1-containing immune complexes, thereby altering quantity and quality of the circulating antigen *in vivo*.

**Fig. 2:**
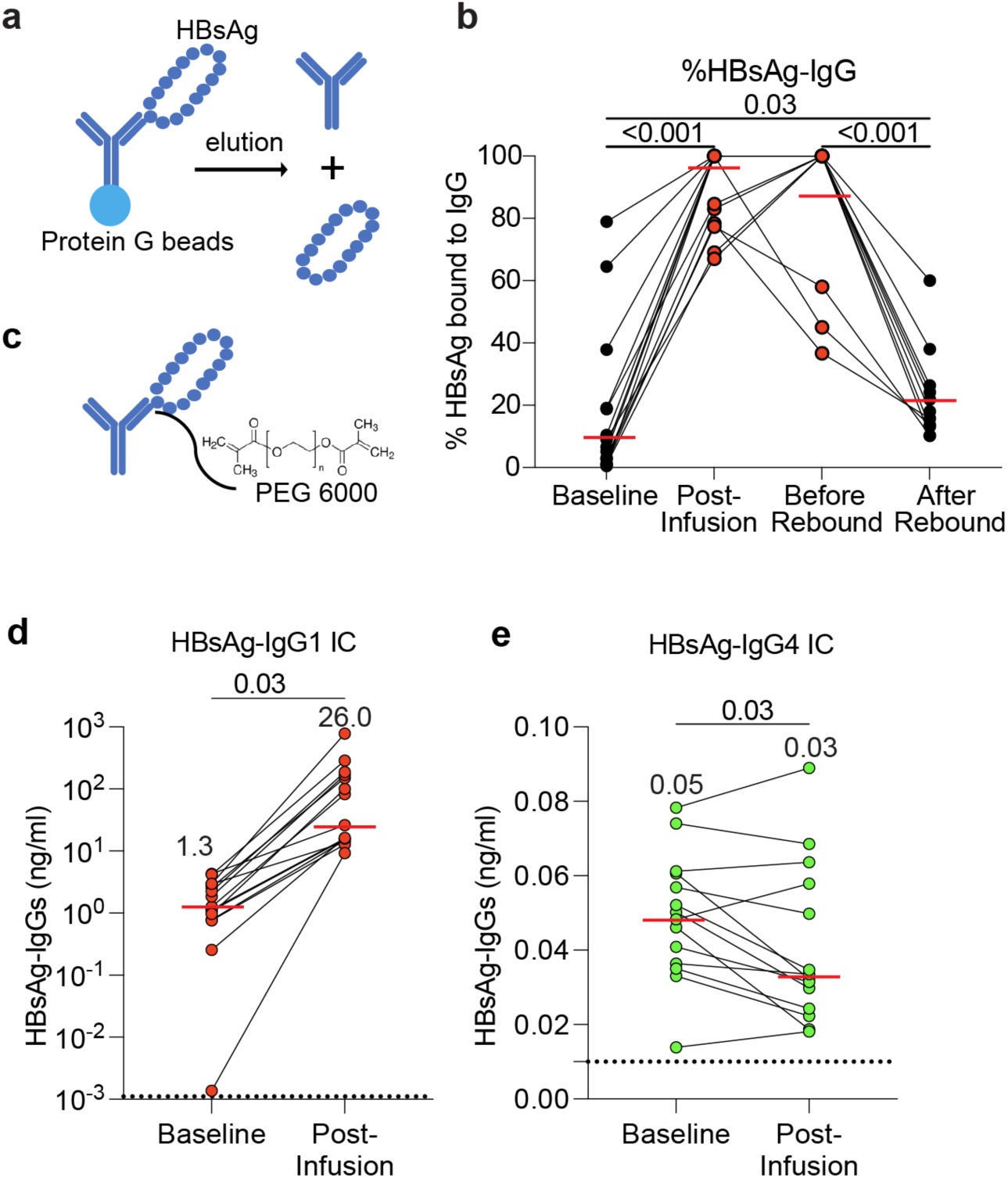
mAb19-LS infusion induces formation of circulating HBsAg–IgG immune complexes dominated by IgG1. **a**, Schematic of the immune complex isolation strategy. Circulating IgG was captured from patient serum using Protein G–conjugated beads. **b**, Percentage of circulating HBsAg bound to IgG at baseline, immediately after mAb19-LS infusion, prior to antigen rebound, and after rebound as determined by ELISA. Each line represents one participant. Red bars indicate mean values 18, 90, 85, 23% at baseline, post-infusion, before and after rebound, respectively. p-values were calculated using paired t-test, n=14. **c**, Schematic of polyethylene glycol (PEG 6000)– precipitation used to isolate circulating HBsAg–IgG immune complexes in **d** and **e**. **d-e**, Quantification of **d** HBsAg–IgG1 **e** HBsAg–IgG4 immune complexes at baseline and 1-day post-infusion by ELISA. Values are shown on a log scale. Each line represents one participant; red bars and numbers indicate median values. Statistical significance in **b**, **d** and **e** was determined using a paired t-test, n=15. Dashed line in **d** and **e** indicates assay lower limit of detection.

Immune complexes between an anti-HBsAg monoclonal antibody with an Fc domain mutation that enhances Fc receptor binding can activate DCs *in vitro* ^23^. To determine whether mAb19- LS immune complexes can mediate Fc receptor activation, we incubated serum obtained from pre- and post-mAb19-LS infusion time points with an IL-2 producing indicator cell line expressing human FcγRIIa and FcγRIIIa ^27,28^. Serum from 6-hour post-infusion showed significantly increased stimulation of IL-2 production compared to baseline for both FcγRIIa and FcγRIIIa (Supplementary Fig. 6). Thus, circulating immune complexes formed shortly after infusion of mAb19-LS can mediate FcγR-IIa and FcγRIIIa activation.

To determine whether circulating antigen presenting cells take up the newly formed immune complexes, we examined permeabilized cells by flow cytometry using fluorochrome-conjugated anti-HBsAg antibodies recognizing epitopes that did not overlap with mAb19 (Fig. 3a-h, Supplementary Fig. 7). Antibody-infused but not placebo-treated individuals showed significant increases in circulating cDC1 and cDC2 associated HBsAg, with a similar trend in monocytes (Fig. 3a-h).

**Fig. 3:**
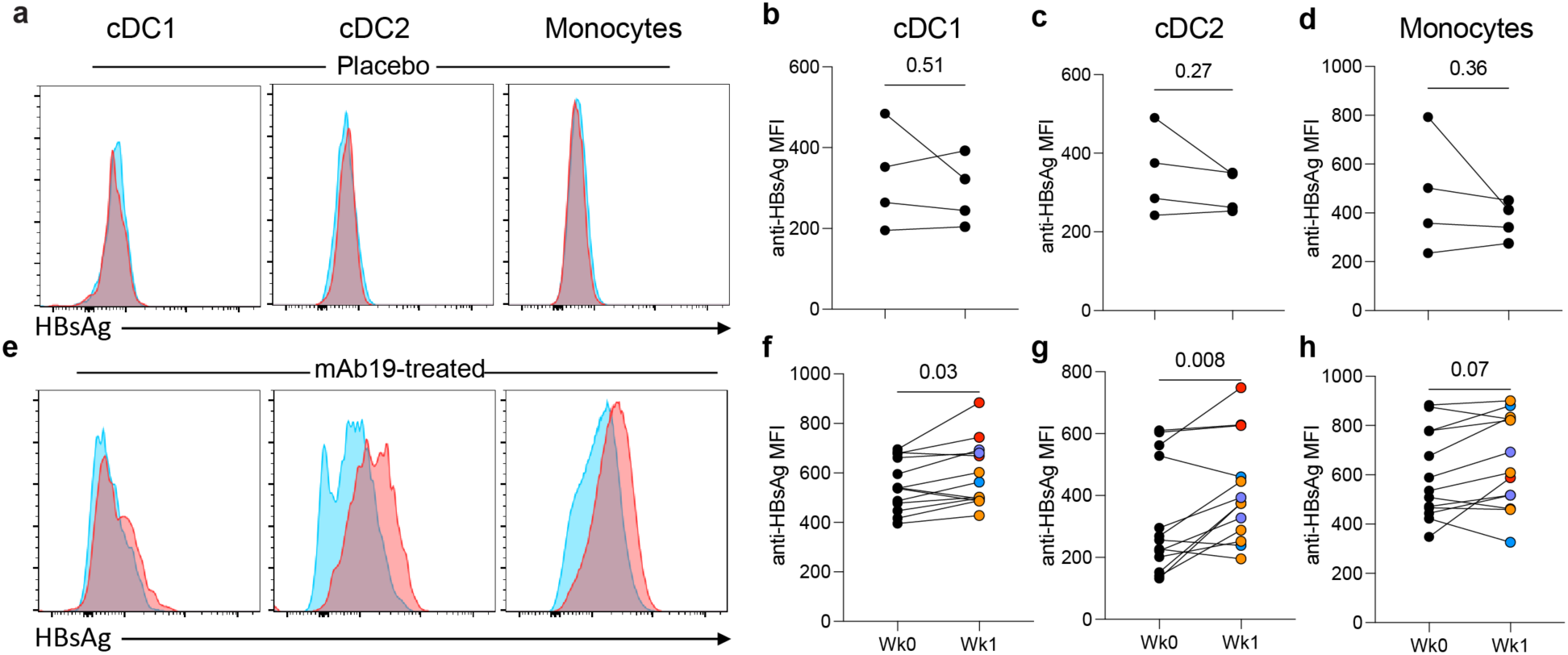
HBsAg-IgG immune complex uptake by circulating myeloid cells. (**A**) Representative flow cytometry histograms showing intracellular HBsAg detection (anti-HBsAg staining) in permeabilized circulating cDC1, cDC2, and monocytes from placebo participants at week 0 (blue) and week 1 (red). (**B–D**) Quantitation of anti-HBsAg mean fluorescence intensity (MFI) on (**B**) cDC1, (**C**) cDC2, and (**D**) monocytes from placebo participants at week 0 (Wk0) and week 1 (Wk1). Each line represents one participant. (**E**) Representative flow cytometry histograms showing HBsAg detection (anti-HBsAg staining) in permeabilized circulating cDC1, cDC2, and monocytes from mAb19-treated participants at week 0 (blue) and week 1 (red). (**F–H**) Quantification of anti-HBsAg MFI on (**F**) cDC1, (**G**) cDC2, and (**H**) monocytes from mAb19-treated participants at week 0 (Wk0) and week 1 (Wk1). Each line represents one participant. Colored symbols denote individual participants, participants that received 1mg/kg (orange), 3mg/kg (blue), 10mg/kg(red) and 30mg/kg (purple). Statistical significance in **B**, **C**, **D**, **F**, **G**, **H** was determined using a paired t-test.

DCs activated with immune complexes *in vitro* can stimulate memory CD4^+^ T cell proliferation and cytokine secretion in chronically infected individuals ^23^. Given the central role of DCs in shaping T cell tolerance and immunity ^29^, we examined whether the mAb19-LS infusion altered T cell responses. To this end, we incubated peripheral blood mononuclear cells (PBMCs) obtained from pre-infusion and week 12 or 24 post-infusion time points with HBV peptides *in vitro* and measured CD4⁺ and CD8⁺ T cell proliferative responses and CD62L expression. CD4⁺ and CD8⁺ T cells obtained at week 12 showed significantly higher levels of proliferation than pre-infusion controls irrespective of CD62L expression (Fig. 4a-f). The magnitude of the proliferative response decreased at week 24 but remained significantly increased when compared to pre-infusion levels for all but CD8^+^CD62L^+^ cells (Fig. 4a-f).

**Fig. 4:**
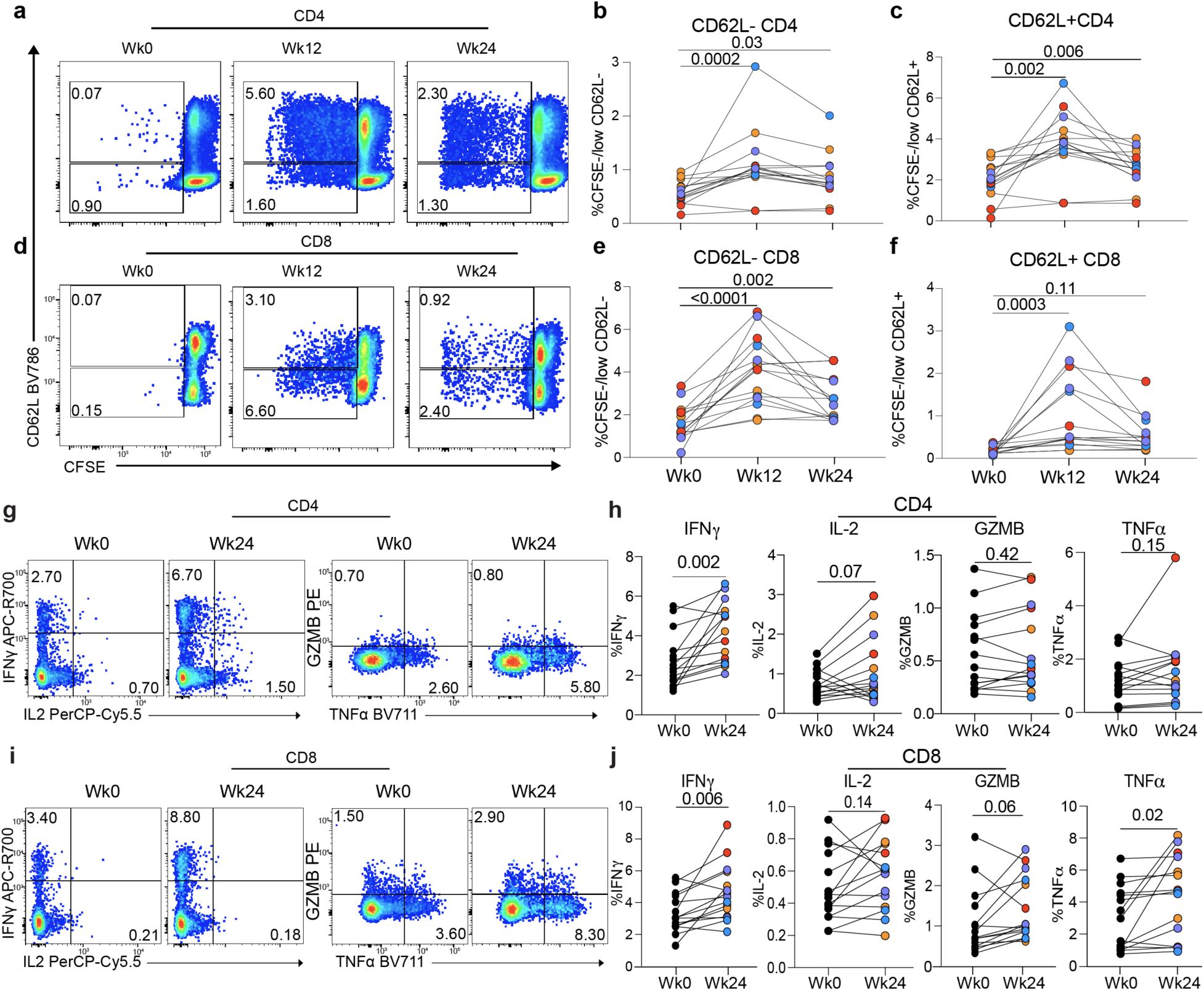
Longitudinal analysis of T cell proliferation and effector cytokine production. **a**, Representative flow cytometry plots showing CFSE dilution versus CD62L expression in CD4⁺ T cells at week 0 (Wk0), week 12 (Wk12), and week 24 (Wk24) after 7 days of *in vitro* stimulation with HBV peptides. Quadrants indicate CFSE^low/-^ CD62L⁻ and CFSE ^low/-^ CD62L⁺ populations. **b-c** Quantitation of proliferating CD4⁺ T cell subsets pre-infusion and 12 or 24 weeks after infusion. **b**, Percentage of CFSE^low/-^ CD62L⁻ cells within CD4⁺ T cells. **c**, Percentage of CFSE^low/-^ CD62L⁺ cells within CD4⁺ T cells. Each line represents one participant. **d**, Representative flow cytometry plots showing CFSE dilution versus CD62L expression in CD8⁺ T cells at Wk0, Wk12, and Wk24 after 7 days of *in vitro* stimulation with HBV peptides. Quadrants indicate CFSE ^low/-^ CD62L⁻ and CFSE ^low/-^ CD62L⁺ populations. **e-f**, Quantitation of proliferating CD8⁺ T cell subsets across timepoints. **e**, Percentage of CFSE^low/-^ CD62L⁻ cells within CD8⁺ T cells. **f**, Percentage of CFSE ^low/-^ CD62L⁺ cells within CD8⁺ T cells. Each line represents one participant. **g**, Representative intracellular cytokine staining plots of CD4⁺ T cells at Wk0 and Wk24 following with HBV peptides stimulation, showing IFN-γ versus IL-2 and GZMB versus TNF-α expression. **h**, Quantitation of cytokine-producing CD4⁺ T cells at Wk0 and Wk24, including frequencies of IFN-γ⁺, IL-2⁺, GZMB⁺ and TNF-α⁺cells. **i**, Representative intracellular cytokine staining plots of CD8⁺ T cells at Wk0 and Wk24 showing IFN-γ versus IL-2 and GZMB versus TNF-α expression. **j**, Quantitation of cytokine-producing CD8⁺ T cells at Wk0 and Wk24, including frequencies of IFN-γ⁺, IL-2⁺, GZMB⁺ and TNF-α⁺cells. Each line represents one participant. Dots represent individual participants. Statistical significance in **b**, **c**, **e** and **f** was determined using a nonparametric one-way ANOVA, and in **h** and **j** using a paired t-test. Colored symbols denote individual participants, participants that received 1mg/kg (orange), 3mg/kg (blue), 10mg/kg (red) and 30mg/kg (purple).

To determine the functional consequences of alterations in T cell responses at week 24 post-infusion, we measured cytokine production by CD4^+^ and CD8^+^ T cells. Compared to baseline, CD4⁺ T cells obtained from week 24 showed significantly increased IFN-γ expression (p=0.002), with a trend toward elevated IL-2 production (Fig. 4g,h). Week 24 CD8⁺ T cells exhibited significant increases in IFN-γ and TNF-α production, along with trends toward enhanced granzyme B and IL-2 production (Fig. 4i,j), indicating enhanced effector activity.

To assess whether immune complex containing antigen presenting cells obtained from mAb19- LS infused individuals would elicit T cell proliferative responses in the absence of exogenously added peptides, autologous T cells were co-cultured with DCs obtained from pre-infusion or 24 weeks post-infusion samples in cross-over assays (Fig. 5a-d). In the absence of additional antigen, proliferative responses were obtained with the combination of CD4^+^ or CD8^+^ T cells and DCs obtained from week 24. Notably, DCs obtained at week 24 elicited greater CD4⁺ and CD8⁺

**Fig. 5:**
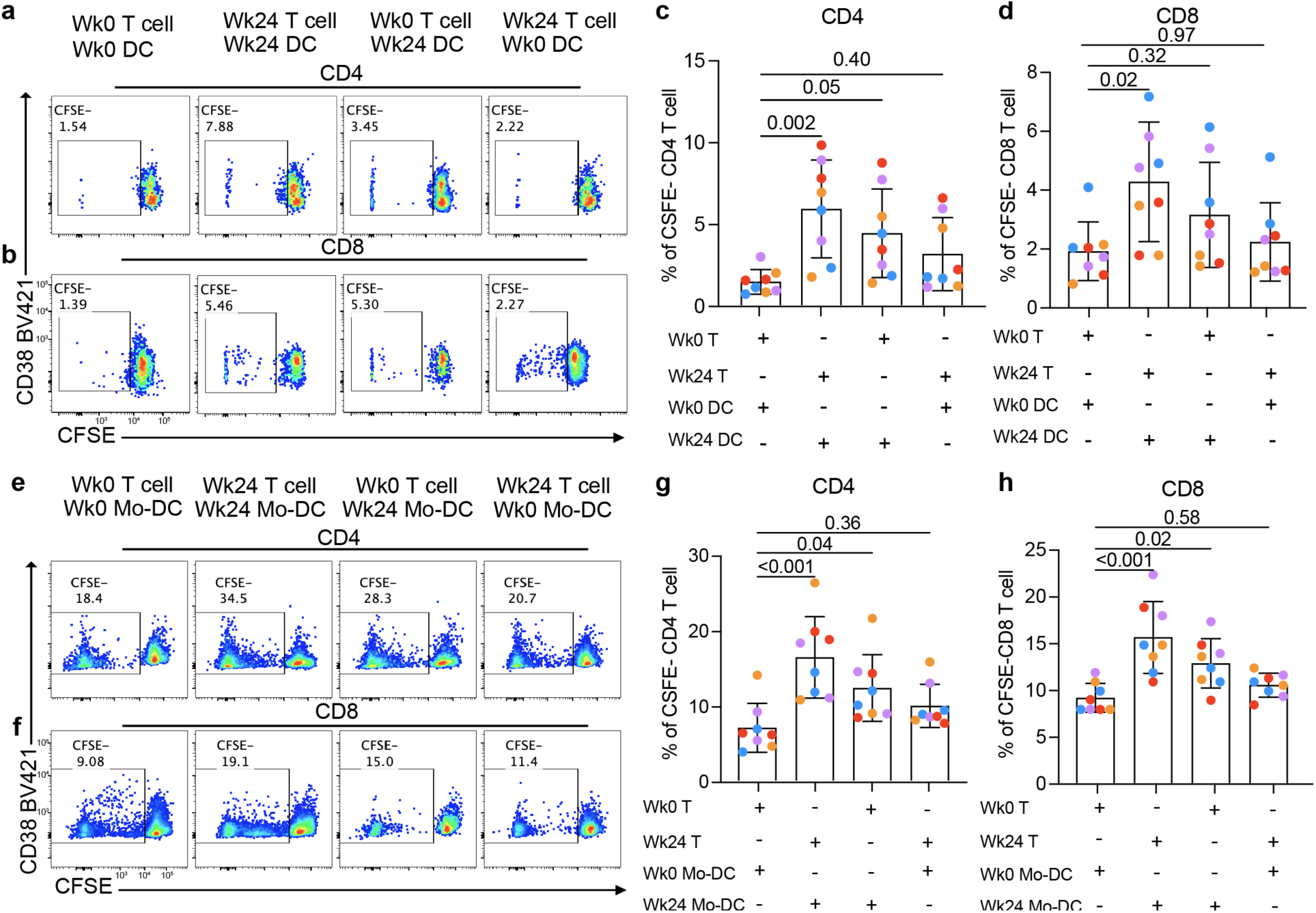
Enhanced T cell proliferation induced by DCs and Mo-DCs after mAb19-LS infusion. **a,b**, Representative flow cytometry plots showing CFSE dilution and CD38 expression in CD4⁺ **a**, and CD8⁺ **b**, T cells following co-culture with DCs isolated at week 0 (Wk0) or week 24 (Wk24). T cells and DCs were collected at the indicated time points, and proliferating cells were identified as the CFSE^low/^⁻ population. **c,d**, Quantitation of the frequency of CFSE ^low/^⁻ CD4⁺ (**C**) and CD8⁺ **d** T cells when cocultured with DCs as indicated in the table. **e,f**, Representative flow cytometry plots showing CFSE dilution and CD38 expression in CD4⁺ **e**, and CD8⁺ **f** T cells following co-culture with monocyte-derived dendritic cells (Mo-DCs) obtained at Wk0 or Wk24. Proliferating cells were identified as the CFSE ^low/^⁻ population. Quantitation of CFSE ^low/^⁻ CD4⁺ **g** andCD8⁺ **h** T cells when cocultured with Mo-DCs as indicated in the table. Bars represent mean ± s.d.; dots represent individual participants. Statistical significance in **c**, **d, g** and **h** was determined using a nonparametric one-way ANOVA. Colored symbols denote individual participants, participants that received 1mg/kg (orange), 3mg/kg (blue), 10mg/kg(red) and 30mg/kg(purple).

T cell proliferative responses than pre-infusion DCs regardless of whether the T cells were obtained from pre-infusion or week 24 time points. In contrast, T cells isolated at week 24 did not display enhanced proliferative responses when stimulated with pre-infusion DCs (Fig. 5a-d). Thus, DCs that take up mAb19-LS HBsAg immune complexes *in vivo* can elicit endogenous CD4^+^ and CD8^+^ T cell proliferative responses in the absence of additional antigen or immune stimulants. Production of monocyte derived DCs requires a 6-day *in vitro* culture with GM-CSF and IL-4 during which ingested immune complexes were degraded. Nevertheless, similar results were observed using monocyte-derived DCs co-cultured with autologous T cells after addition of HBV peptides (Fig. 5e-h). These findings suggest that mAb19-LS-HBsAg immune complex associated changes in antigen presenting cells contribute to enhanced T cell activation.

To determine whether there were transcriptional changes corresponding to results of the co-culture experiments, we performed single cell RNA sequencing (scRNA-seq) on 3 treated and 1 placebo participant using samples collected pre-infusion and at week 24, a time when the treated individuals remained suppressed. CD4^+^ and CD8^+^ T cells were enriched by cell sorting and processed for 10X Genomics sequencing. Cells were distributed across 10 distinct clusters corresponding to subsets of CD4+ and CD8+ T cells, as visualized by uniform manifold approximation and projection (UMAP) (Supplementary Fig. 8a). Overall T cell subset composition was largely preserved after infusion. However, consistent with enhanced CD8⁺ T cell effector activity observed in antigen-stimulation assays (Fig. 4k-n), the frequency of CD8⁺ effector memory T cells increased at week 24 in treated participants (Supplementary Fig. 8b). Differential expression analysis identified a limited set of activation-associated transcripts increased at week 24, including *DUSP2* and *PDE4B* in both CD4^+^ and CD8^+^ T cells (Supplementary Fig. 8c-d). Gene set enrichment analysis (GSEA) of the pooled data from treated participants revealed significant enrichment of inflammatory programs at week 24, including TNF-α signaling via NF-κB in both CD4^+^ and CD8^+^ T cells and IFN-γ signaling in CD4^+^ T cells, with a similar trend in CD8^+^ T cells (Fig. 6a). Analysis at the individual participant level showed that these pathway changes were heterogeneous in magnitude, but TNF-α signaling, mTORC1 signaling, IFN-γ response, and inflammatory response signatures were generally increased in mAb19-LS recipients (Fig. 6b).

**Fig. 6:**
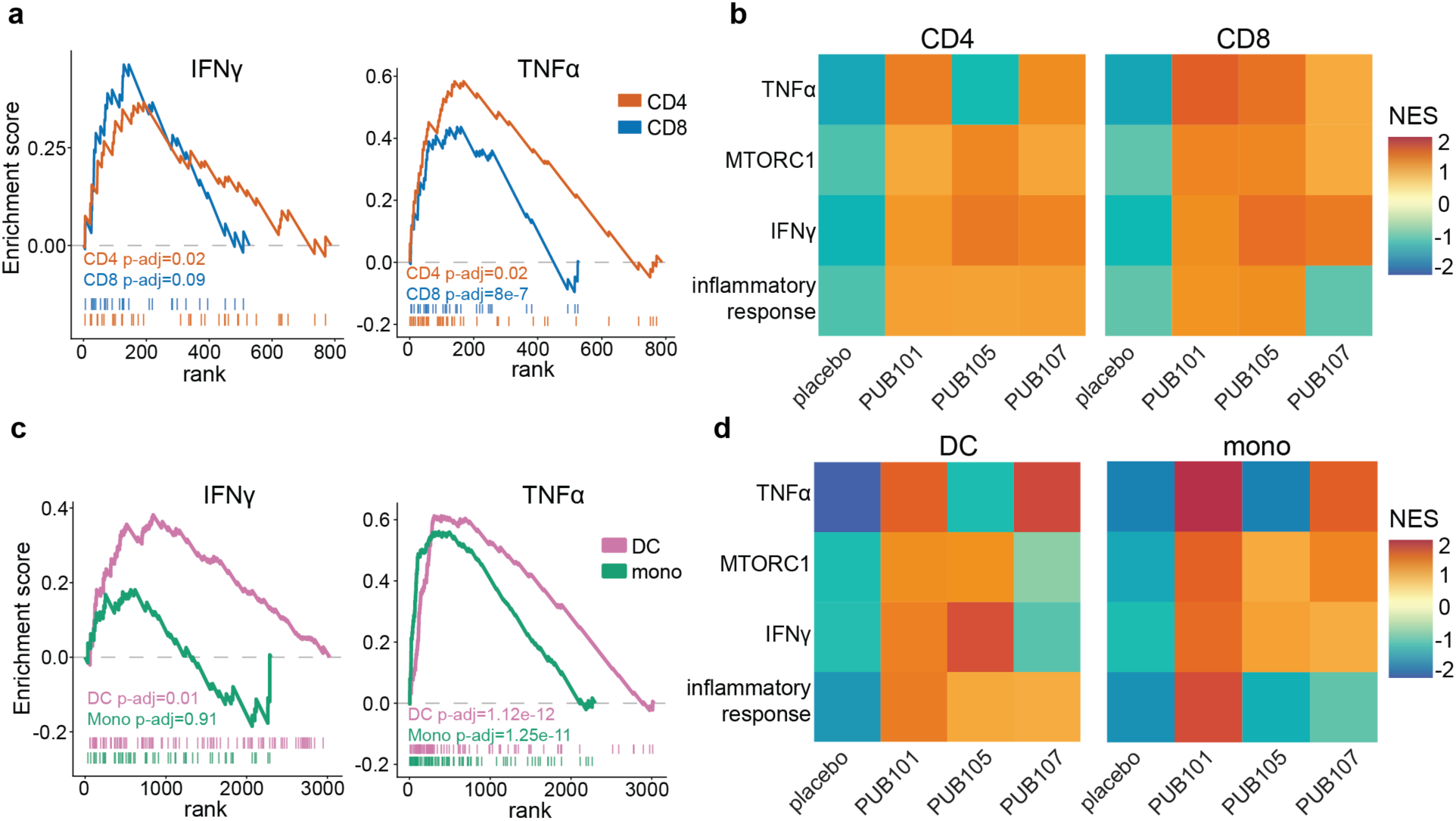
Single-cell transcriptomic analysis of T cells, cDCs and monocytes. **a**, Gene set enrichment analysis (GSEA) comparing week 24 and baseline CD4⁺ T cell and CD8⁺ T cell transcriptomes from 3 combined mAb19-LS recipients (see methods). GSEA plots are shown for IFN-γ signaling (left) and TNF-α signaling via NF-κB pathways (right). Curves represent the enrichment score, and corresponding vertical bars indicate gene positions within the ranked gene list. IFN-γ: CD4 p-adj=0.02, CD8 p-adj=0.09; TNF-α: CD4 p-adj=0.02, CD8 p- adj=8e-7. **b**, Individual participant GSEA analysis of CD4⁺ and CD8⁺ T cells from mAb19-LS recipients and placebo at Wk24 compared to baseline. Heat maps show normalized enrichment scores (NES) for TNF-α signaling, mTORC1 signaling, IFN-γ response and inflammatory response signatures for CD4^+^ (left) and CD8^+^ (right) T cells. **c**, GSEA comparing baseline and week 24 DC and monocyte transcriptomes from combined mAb19-LS recipients. GSEA plots are shown for IFN-γ signaling (left) and TNF-α signaling via NF-κB pathways (right). Curves represent enrichment score, and corresponding vertical bars indicate gene positions within the ranked gene list. IFN-γ: DC p-adj=0.013, monocyte p-adj=0.91; TNF-α: DC p-adj=1.12e-12, monocyte p-adj=1.25e-11. **d**, Individual participant pathway analysis of DCs (left) and monocytes (right) from mAb19-LS recipients and placebo at Wk24 compared to baseline. Heat maps show NES for TNF-α signaling, mTORC1 signaling, IFN-γ response and inflammatory response signatures.

To determine whether enhanced antigen uptake and presentation by DCs and monocytes was accompanied by phenotypic changes in these cells, we performed scRNA-seq on DCs and monocytes sorted from matched samples from the same participants and time points used for the T cell analysis. Cells clustered into 6 unique DC/monocyte subsets visualized using UMAP (Supplementary Fig. 8e) with no significant changes in distribution at week 24 compared to baseline (Supplementary Fig. S8f). In contrast to the relatively modest changes observed in T cells, DCs and monocytes showed sustained inflammatory changes in genes involved in IFN-γ and TNF-α signaling pathways (Supplementary Fig. 8g,h). DCs differentially expressed genes 24 weeks after infusion, including activation and inflammatory response associated genes such as *NR4A3*, *CDKN1A*, *NR4A1*, *IER3*, *PTGS2* and *DUSP2*, whereas monocytes showed induction of immediate-early and inflammatory response genes including *IER3*, *DUSP2*, *NR4A3*, *CXCL8*, *OSM* and *PTGS2* (Supplementary Fig. 8g,h). GSEA revealed significant upregulation of inflammatory signaling programs, including TNF-α signaling via NF-κB in both DCs and monocytes and IFN-γ–associated pathways in DCs (Fig. 6c). Individual participant analysis similarly showed persistent but variable enrichment of TNF-α signaling, mTORC1 signaling, IFN-γ response, and inflammatory response programs in DCs and monocytes (Fig. 6d). Although the magnitude of these changes varied between individuals, they are indicative of prolonged innate immune reprogramming following mAb19-LS therapy.

## Discussion

Persistent circulating HBsAg contributes to profound immune dysfunction in chronic HBV infection. IFN-α and suppressive siRNAs are reported to enable sustained remission ^30–32^. However, remission is limited to 20-30% of individuals with initial HBsAg levels below 1000 IU/ml, and 10% or less of those with HBsAg levels 1,000-3,000 IU/ml ^30–32^. How antiviral immunity might be further enhanced in humans remains unclear—particularly because HBV-specific T cell dysfunction reflects heterogeneous, liver-imposed differentiation constraints that do not fully conform to classical terminal exhaustion ^11^.

Monoclonal antibodies are important therapeutics in cancer and autoimmune disease that enhance immune function by specific antigen binding and subsequent engagement of innate and adaptive immune cells ^33^. Our study demonstrates that a single infusion of anti-HBsAg IgG1 antibody mAb19-LS was generally well tolerated at doses ranging from 1 to 30 mg/kg. Transient grade 1 or 2 infusion reactions occurred in 20% of participants (3 of 15), with associated short-lived lymphopenia, as well as grade 2 elevation in transaminases in one participant. These effects occurred in participants with higher baseline HBsAg levels and was not associated with mAb19-LS dose level.

mAb19-LS rapidly converted nearly all freely circulating HBsAg into IgG1-bound immune complexes. The result was accelerated antigen clearance and HBsAg uptake by antigen presenting cells. These findings are consistent with biochemically documented properties of the Fc domain of human IgG1 which binds to nearly all Fc receptors, with particularly high affinity to activating Fcg receptors found on DCs and monocytes ^33^.

In the steady state, DCs help maintain immune tolerance by presenting self-antigens found in circulation and tissues to T cells under conditions that prevent their differentiation into effector cells^29,34,35^. In chronic HBV infection the preponderance of the circulating antigen enters DCs by endocytosis without inducing DC activation ^36,37^. However, *in vitro* experiments showed that Tobevibart (VIR-3434), an Fc enhanced anti-HBsAg antibody, can form immune complexes capable of activating antigen presenting cells ^23^. mAb19-LS infusion produced large quantities of IgG1-HBsAg immune complexes that induced DC differentiation to a more inflammatory and immune stimulatory state *in vivo*, enabling them to activate T cells to produce IFN-γ and TNF-α, effector cytokines associated with HBV clearance ^5–7,38,39^. Together, these findings suggest that antibody therapy is associated with measurable remodeling of antiviral immune programs in chronic HBV infection.

These results are notable in the context of recent mechanistic advances, indicating that HBV-specific CD8⁺ T cells occupy heterogeneous dysfunctional states shaped by antigen presentation in the liver ^11^. In mouse models, intrahepatic priming can imprint dysfunctional programs distinct from canonical exhaustion ^11^. More recently, single-cell and functional analyses have resolved distinct dysfunctional subsets induced by hepatocellular priming that are relatively refractory to inhibitory receptor blockade, but can be redirected by co-stimulatory TNFR agonism, particularly 4-1BB ^15^. Complementing these findings in humans is an “attenuated cytotoxic” HBV-specific CD8⁺ T cell subset associated with viral control that is regulated by TGFβ signaling, highlighting that constrained antiviral activity is reversible ^12,16,40^. Our data extend this emerging framework by providing evidence in chronically HBV infected individuals that immune complexes produced by mAb19-LS infusion can enhance antigen presentation and T cell responses.

Clinical efforts to achieve remission through HBsAg suppression provide an important benchmark. Recent phase 2 and 3 studies with combinations of siRNA and/or pegylated interferon and antisense oligonucleotides show that persistent HBsAg loss is achievable in up to 20-30% of treated individuals with low levels of circulating HBsAg ^30–32,41^, reinforcing the promise—and complexity—of combining antigen suppression with immune modulation. Creation of immune complexes by antibody therapy is mechanistically distinct from both transcriptional silencing by siRNA and pegylated interferon therapy. The key difference is that Fc engagement by mAb19-LS immune complexes alters antigen handling *in vivo* and subsequent DC-T cell interactions in ways that favor effector T cell responses. Our results are further supported by experiments with Fc-engineered anti-HBsAg antibody Tobevibart, (VIR-3434) that showed Fc receptor–mediated HBsAg uptake by antigen presenting cells *in vitro* ^23^.

This study has limitations. Cohort sizes were modest and tissue sampling was not available, precluding direct assessment of intrahepatic events that likely shape HBV-specific T cell fate. Single-cell profiling was performed in a limited number of participants, and the trial was not powered to evaluate long-term clinical endpoints. Future studies with larger cohorts, longer follow up, repeat dosing, deeper longitudinal sampling, and integration with intrahepatic sampling where feasible will be required to define durability, mechanisms, and clinical relevance.

Finally, our findings support the view that universal durable immune control of chronic HBV will likely require combination strategies. Recent mechanistic work suggests that redirecting dysfunctional HBV-specific CD8⁺ T cells may require defined co-stimulatory or cytokine-mediated interventions. Antibody-based therapies offer complementary advantages—including long half-life, immune complex formation, and Fc-dependent immunomodulation—that could be rational partners for antiviral suppression and targeted immune activators. In sum, our study indicates that anti-HBsAg immunotherapy is generally safe and links antibody-mediated antigen modulation to coordinated remodeling of antiviral immune responses in humans, supporting further clinical evaluation of immune complex–based strategies within multi-modal approaches toward functional cure.

## Data Availability

All data produced in the present work are contained in the manuscript.

## Acknowledgments

We thank members of the Nussenzweig lab for discussions and K. Gordon for flow cytometry support. We acknowledge the use of the Integrated Genomics Core. CG is a Charité- Foundation Recruiting Grantee and was supported by the HJH-Foundation. MCN is a Howard Hughes Medical Institute investigator (HHMI). This article is subject to HHMI’s Open Access to Publications policy. HHMI lab heads have previously granted a non-exclusive CC BY 4.0 license to the public and a sublicensable license to HHMI in their research articles. Pursuant to those licenses, the author-accepted manuscript of this article can be made freely available under a CC BY 4.0 license immediately upon publication.

## Author Contributions

Experimental design/execution: ZW, MT, HNL, SNHS, GB, AKJ, MS, CG, OSS, MC, MCN

Trial physicians: HNL, SNHS, JDG, VI, HG, CZ, MK, NW, RS, PTL, DH, HY, CP, ISJ

Sample collection/processing: AKJ, MS, VV, JD, IS, DD

Protocol writing/study design: SDWA, TR, MC

Recruitment: HNL, SNHS, CZ, KM, MK, AMN, MD, MC, FF, DD, KM

Institutional lead/co-PI: FT, LES, FK

Mathematical modeling: DBR

Bioinformatics: GSSS

Antibody characterization/production: ZW, QW, BH, AG

Supervision: ZW, CG, OSS, MC, MCN

Writing – original draft: ZW, MT, MCN

Writing – editing and reviewing: all authors

## Data Availability

Transcriptomic data generated in this study are available under GSA accession ### (made available upon publication).

## Methods

### Study design

Two parallel placebo-controlled dose-escalation phase I studies were conducted in adults with chronic HBV infection receiving suppressive nucleos(t)ide analogue therapy (NCT05856890, EUCT number: 2023-508444-22-00 and NCT06668727). The two studies have identical study designs and scheduled assessments, and similar eligibility criteria (see below). Participants were randomized 3:1 to receive mAb19-LS or placebo (sterile saline). mAb19-LS was administered as a single intravenous infusion at dose levels of 1 mg/kg, 3 mg/kg, 10 mg/kg, or 30 mg/kg. All participants provided written informed consent prior to enrollment. The study was conducted in accordance with Good Clinical Practice guidelines and approved by the Institutional Review Boards of the Rockefeller University in the United States, and the National Regulatory and Ethics Committees in Denmark and Germany under EUCT number: 2023- 508444-22-00.

### Study drug

mAb19-LS is a fully human IgG1 monoclonal antibody recognizing hepatitis B surface antigen (HBsAg). mAb19 was cloned from an individual immunized with an approved standard HBsAg^25^. The LS mutation (M428L/N434S) in the Fc–FcRn interaction domain of IgG1 enhances FcRn binding and intracellular recycling, increasing antibody half-life ^42^. mAb19-LS was expressed in Chinese hamster ovary (CHO) cells and purified by protein A chromatography under Good Manufacturing Practice (GMP) conditions by Celldex Therapeutics. The drug substance was formulated at 20 mg ml⁻¹ in phosphate-buffered saline containing 0.01% polysorbate 80 and supplied as single-use sterile vials for intravenous administration. Vials were stored at ≤ -65°C until use.

### Study participants

Eligible participants were adults aged 18–70 years with documented chronic HBV infection who had received stable antiviral therapy for at least 12 months before enrollment, had quantifiable serum HBsAg levels, and had HBV DNA levels below the limit of quantification. The European study excluded HBeAg positive participants, while the US study did not exclude based on HBeAg status. Main exclusion criteria in both studies included: cirrhosis or grade 3 or higher liver fibrosis, co-infection with hepatitis D or C virus or HIV, history of chronic liver disease of another cause, history of immunosuppression in the last 6 months, prior receipt of anti-HBV mAb, and clinically significant laboratory abnormalities, and pregnancy. Baseline demographic and clinical characteristics are provided in supplementary table 1.

### Study procedures

The appropriate dose of mAb19-LS was calculated based on body weight and diluted in sterile normal saline to a total infusion volume of 100–500 ml. Placebo recipients received the equivalent volume of sterile saline. The infusion of mAb19-LS or placebo was administered intravenously over 60 minutes. Participants were monitored for 6 hours following infusion and returned for scheduled follow-up visits at 1, 3 and 7 days after mAb19-LS or placebo infusion, weekly through week 4, and at weeks 8, 12, 24, 36 and 48. All participants have completed at least 48 weeks of follow up. At each visit, safety assessments for the occurrence of adverse events and physical examination were performed. Serum HBsAg levels were measured at each visit, and other laboratory tests (including plasma HBV DNA, complete blood cell counts, chemistries, liver function tests, coagulation times and completement factors C3 and C4) were performed at selected visits. Blood samples (30–120 ml) were collected prior to infusion and at multiple time points post-infusion. Samples were processed within 4 hours of collection. Serum and plasma were stored at –80 °C. PBMCs were isolated by density gradient centrifugation and cryopreserved in fetal bovine serum with 10% DMSO.

### Measurement of mAb19-LS serum levels

Serum concentrations of mAb19-LS were determined using a validated anti-idiotype sandwich ELISA. High-binding 384-well plates were coated overnight at 4°C with 2 µg ml⁻¹ of an anti-idiotypic monoclonal antibody specific for the variable region of mAb19. Plates were blocked with 2% BSA. Serum samples were diluted (minimum 1:100) and incubated for 1 hour at 37°C. Bound mAb19-LS was detected using HRP-conjugated anti-human IgG Fc antibody and developed with tetramethylbenzidine substrate. Concentrations were interpolated from a standard curve generated using purified mAb19-LS drug product and fitted with a four-parameter logistic model. Specificity was confirmed using pre-dose serum and unrelated human IgG1 controls.

### Detection of circulating HBsAg–IgG immune complexes in serum

Circulating immune complexes were quantified using a Protein G capture assay followed by HBsAg ELISA detection ^43^. Serum samples were diluted 1:10 in PBS and added to Protein G Sepharose beads (Cytiva #17061806) and incubated overnight at 4°C, rotating. After washing the beads with PBS, captured IgG and associated immune complexes were eluted using IgG elution buffer (Thermo Scientific #21009), neutralized, and measure by ELISA. HBsAg was assessed by chemiluminescence using HBsAg CLIA kit (DiaSino #DS187701). In selected experiments, immune complexes were also enriched by polyethylene glycol (PEG) precipitation ^44^. Serum was mixed with PEG 6000 to a final concentration of 12.5% and incubated overnight at 4°C. Samples were then centrifuged at 10,000g for 30 min, and the resulting pellets were resuspended in PBS. Resuspended precipitates were analyzed by ELISA to determine immune complex IgG subclass distribution.

### FcR activation by circulating HBsAg–IgG immune complexes *in vitro*

FcγR activation was measured using a previously described assay^28^. In brief, 6×10^4^ mouse BW5147 reporter cells stably expressing FcγRIIA or FcγRIIIA were mixed with two-fold serial dilutions of human serum in a total volume of 30 µl. Serial dilutions of HepB mAb19 were used as negative control. Mixtures were incubated for 17 h at 37°C and 5% CO2 in 384-well plates pre-treated with PBS/10% FCS overnight at 4°C.

Mouse IL-2 expression was quantified by ELISA as described earlier ^23^. Briefly, cell supernatants were diluted 1/2 in PBS with 5% BSA and 0.1% Tween-20, and mIL-2 was measured using the capture antibody JES6-1A12 and the biotinylated detection antibody JES6- 5H4 ^23^.

### Flow cytometry

PBMCs were isolated by Ficoll density centrifugation and cryopreserved. Thawed cells were rested for 2 hours in 5% CO2 incubator at 37°C prior to staining. PBMCs were first stained for surface markers (CD3, CD4, CD8, CD14, CD16, CD19, CD335, BDCA-1, BDCA-2, BDCA-3, HLA-DR) for 20 minutes at 4°C. Cells were then fixed using 4% paraformaldehyde for 15 minutes at room temperature and permeabilized using BD Cytofix/Cytoperm buffer according to the manufacturer’s instructions. Intracellular HBsAg was detected using a directly conjugated anti-HBsAg monoclonal antibody that recognizes a non-overlapping epitope on HBsAg to mAb19-LS (clone HB017 or HB020^25^). When using unconjugated primary antibody, staining was followed by fluorochrome-conjugated anti-human IgG secondary antibody. Fluorescence-minus-one (FMO) controls and HBV-negative donor PBMCs were used to define gating thresholds. Data were acquired on BD FACSymphony A5 Cell Analyzer and analyzed in FlowJo v10.

### Intracellular cytokine staining (ICS)

Cryopreserved PBMCs were thawed, washed, and rested for 2 hours at 37°C in RPMI 1640 supplemented with 10% fetal bovine serum and 1% penicillin–streptomycin. For antigen-specific stimulation, PBMCs were incubated with overlapping peptide pools spanning HBsAg and HBcAg (2 µg ml⁻¹ per peptide; JPT Peptide Technologies). For polyclonal stimulation controls, cells were stimulated with phorbol 12-myristate 13-acetate (PMA, 50 ng ml⁻¹) and ionomycin (1 µg ml⁻¹). Brefeldin A (10 µg ml⁻¹) was added during the final 4 hours of stimulation. Total stimulation time was 12 hours at 37 °C. Cells were first stained for viability and surface markers (CD3, CD4, CD8, PD-1, TIM-3, CXCR5, CD45RA, CCR7). Following fixation and permeabilization using BD Cytofix/Cytoperm buffer, intracellular staining was performed for IFN-γ, TNF-α, IL-2, and granzyme B. Data were acquired on FACSymphony A5 Analyzer and analyzed using FlowJo v10.

### CFSE proliferation assay

To assess T cell proliferative capacity, PBMCs were labeled with carboxyfluorescein diacetate succinimidyl ester (CFSE; Thermo Fisher Scientific) at 2.5 µM for 10 minutes at 37°C. The reaction was quenched with complete medium, and cells were washed twice prior to culture. CFSE-labeled PBMCs were cultured in 96-well round-bottom plates at 1 × 10⁶ cells ml⁻¹ and stimulated with HBsAg peptide pools (2 µg ml⁻¹ per peptide) or anti-CD3/CD28 beads (positive control) for 7 days at 37 °C. At day 7, cells were harvested and stained for surface markers (CD3, CD4, CD8). CFSE dilution was analyzed by flow cytometry. Proliferating cells were identified as CFSE^low/-^ populations within CD4⁺ or CD8⁺ T cell gates.

### DC–T Cell Co-culture

Primary DC subsets were isolated from freshly thawed PBMCs by fluorescence-activated cell sorting. BDCA-1⁺ (CD1c⁺) conventional DC2, BDCA-3⁺ (CD141⁺) conventional DC1, and BDCA- 2^+^ plasmacytoid DC populations were identified within live, lineage-negative (CD3⁻CD19⁻CD56⁻) HLA-DR⁺ cells and sorted to >95% purity using a FACSymphony S6 (BD Biosciences). Sorted DCs were directly co-cultured with autologous CD3⁺ T cells at a 1:10 DC:T ratio without addition of exogenous HBV peptides. In parallel, CD14⁺ monocytes were sorted from PBMCs and cultured for 7 days in RPMI 1640 supplemented with 10% fetal bovine serum, granulocyte–macrophage colony-stimulating factor (GM-CSF) (50 ng/ml), interleukin-4 (20 ng/ml), IL-1β(10 ng/ml), IL-6(1000 IU/ml); TNF-α (20 ng/ml) and PGE2 (1ug/ml) to generate monocyte-derived DCs. Mo-DCs were pulsed with overlapping HBV peptide pools prior to co-culture with autologous T cells at the same ratio. After 4–5 days, T cell activation and function were assessed by CD69 and CD25 expression, intracellular cytokine staining following restimulation, and proliferation by CFSE dilution. After washing, Mo-DCs were co-cultured with autologous CD3⁺ T cells at a 1:4 DC:T ratio for 7 days. T cell activation and function were assessed by expression of CD69 and CD38, intracellular cytokine staining following peptide restimulation, and proliferation by CFSE dilution.

### Single-cell RNA-Seq processing

CD4⁺ and CD8^+^ T cells, cDCs and monocytes were processed using the Chromium Single Cell 5′ platform (10x Genomics) according to the manufacturer’s protocol. Gene expression libraries were prepared using the Chromium Single Cell 5′ Library & Gel Bead Kit v2 and sequenced on an Illumina NovaSeq 6000 platform. scRNA-Seq and Hashtag-oligos unique molecular identifier quantification were performed with Cell Ranger multi v.8.0.1 (10x Genomics) using the GEX reference GRCh38 and analyzed in R with Seurat v.5.1.0. Cells were demultiplexed with MULTISeqDemux, and those classified as doublets or with >10% mitochondrial content and feature counts less than 200 or greater than 5,000 were excluded. Ig or TR genes were regressed out. Sample batches were then merged, scaled and normalized with SCTransform. Individual participant datasets were integrated using Integrate Layers with the anchor-based CCA integration method. Based on their gene expression profile, single cells were visualized in a lower dimensional space using Uniform Manifold Approximation and Projection (UMAP) clustering by selecting the first ten principal components. Subsequently, query cells were mapped onto the PBMC reference-defined cell states^45^ via anchor-based label transfer, whereby cell type identities, including Monocytes, DC, CD4^+^ T and CD8^+^ T cells subtypes, were assigned as the reference label with the highest weighted prediction score, captured as predicted.celltype.l1.

Differential gene expression analysis between Wk24 and Wk0 was performed using FindMarkers. For combined mAb treated participants, we applied the MAST hurdle model, with participant identity and total UMI counts per cell included as latent variables. For comparisons within individual patients, we used the Wilcoxon Rank Sum test. Genes with absolute log2 fold change ≥ 0.5 and p-adj < 0.05 were considered differentially expressed. Genes were subsequently ranked by sign(avg_log2_FC) * -log10 p-value, and gene set enrichment analysis(GSEA)^46^ was performed with default parameters using H: Hallmark gene sets from the Molecular Signatures database (MSigDB)^47^.

### Modeling of HBsAg dynamics

To estimate quantitative attributes of mAb-mediated HBsAg clearance, we used a mechanistic viral dynamics model in a population nonlinear mixed effects (pNLME) modeling framework to simultaneously describe the kinetics of the mAb19-LS and HBsAg. To incorporate the influence of time-varying mAb levels on HBsAg kinetics, we included observed mAb concentrations as a linearly interpolated time-varying regressor term in each model of HBsAg kinetics. (We also used an empirical PK fitting scheme as an alternative to linear interpolation and found similar results). Because longitudinal variability was small in the placebo group, we estimated a small proportional error of *a*=0.1 IU/mL and kept this consistent throughout all model fitting. Given the concurrent antiviral treatment setting, we assumed a simplified model in which HBsAg is produced from a single compartment and did not separately model cccDNA, integrated DNA, or infected-cell proliferation in this setting.

We began with a simple model with constant production and natural clearance of HBsAg which would naturally assume a steady state model for HBsAg *S*(*t*) level in advance of therapy

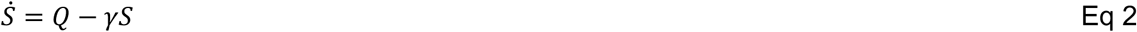

Where the overdot denotes time derivative. This assumes a constant production rate *Q*, which reflects the combined effects of per-cell production and the number of producing cells, and a natural clearance rate *γ*. We therefore can calculate the pre-treatment equilibrium *S*(0) = *Q*/*γ* which can be different for each participant. We then tested models for the action of the mAb, the simplest being a linear clearance with an additional negative term proportional to mAb and HBsAg concentration

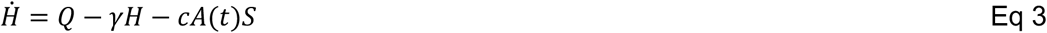

with the mAb induced clearance rate *c* with units [per µg/mL per day]. We also assessed another model with a saturating clearance term

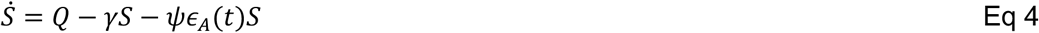

where mAb effect could range from 0-1 relative to concentration following *ε_A_*(*t*) = 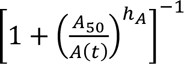. This version fit to data better than the non-saturating model (difference in Bayesian information criterion ΔBIC>100). In this model, mAb-induced clearance follows rate *ψ* with units and two additional parameters are introduced, the concentration *A*_50_ at which clearance is 50% of maximal and the Hill slope ℎ*_A_* that governs how rapidly this function saturates once the concentration shifts above and below the 50% of maximum threshold. We assumed an a priori population parameter distribution for *γ* using existing estimates of HBsAg half-lives of 1.3 95% CI: [0.9–1.8] days and based on sharp transitions to saturation, fixed ℎ*_A_*=10 ^26^. We then estimated population parameters and random effects for *Q*, *A*_50_, and *ψ*, including a correlation between *Q* and *γ*. Residual errors of estimation RSE were all <50% and estimated population values were *Q*=752 IU/mL per day, *γ*=0.37 per day, *ψ*=4.6 per day, and *A*_50_=18.5 µg/mL.

HBsAg half-life was calculated from instantaneous clearance rates as ln 2/ [*γ* + *ψε_A_*(*t*)] and tracked longitudinally. A new equilibrium state was identified for each participant as ≥2 consecutive timepoints where HBsAg had dropped >0.5 log10 from baseline and was changing <0.2 log10 between timepoints; half-life was estimated at the midpoint of this window *t_eq_*.

### Statistical analysis

Statistical analyses were performed using GraphPad Prism v9 and R (v4.4.0) with Rstudio server (2024.04.0 Build 735). For paired comparisons across time points (baseline vs. week 12 or week 24), two-tailed Wilcoxon signed-rank tests were used. For multiple-group comparisons, Kruskal–Wallis tests with Dunn’s correction were applied. Correlation analyses were performed using Spearman’s rank correlation coefficient. P values <0.05 were considered statistically significant. Exact p-values are provided in the figures. No statistical methods were used to predetermine sample size. Investigators were not blinded to group allocation during analysis.

## Supplementary

**Supplementary Fig. 1:**
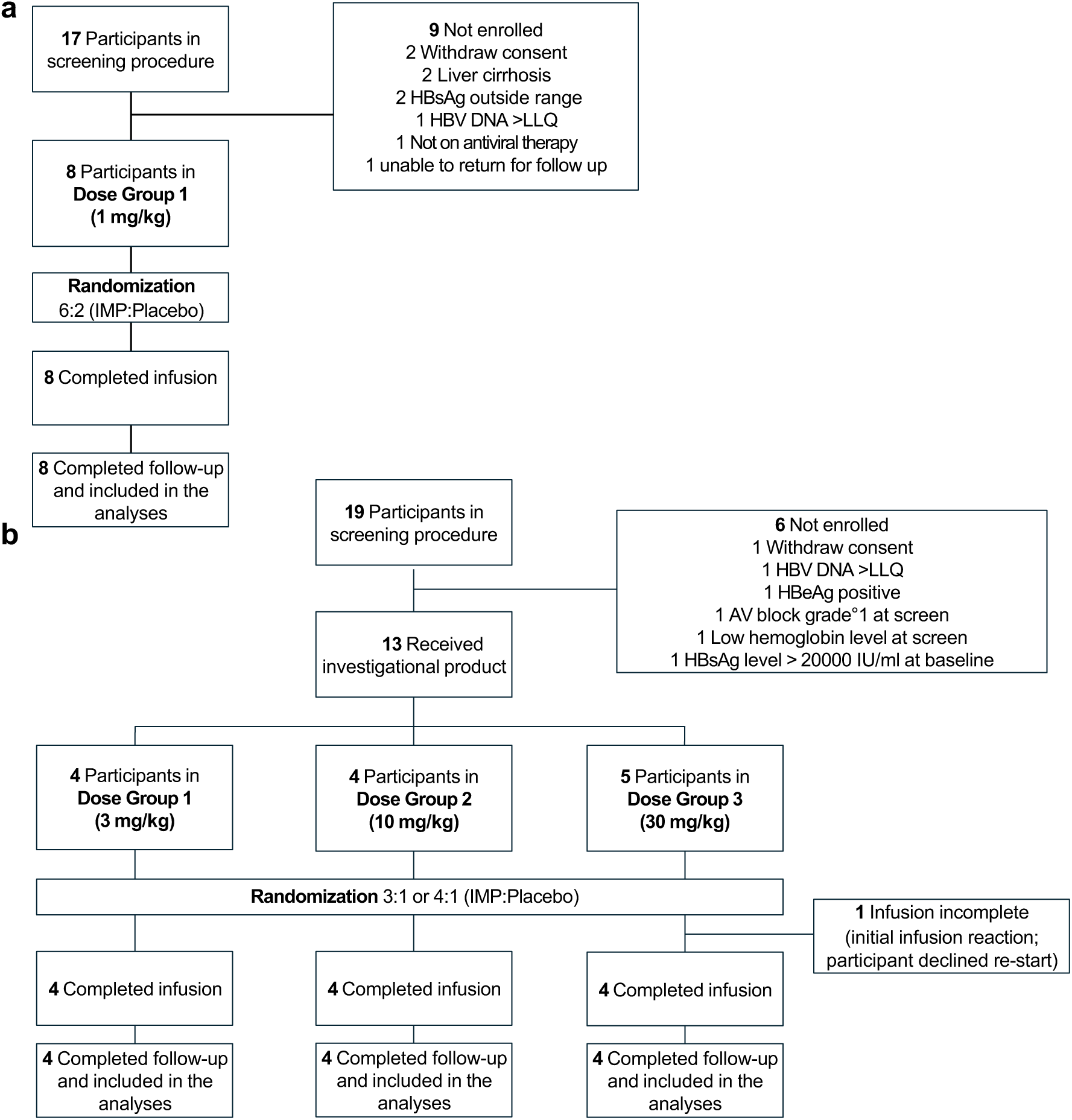
CONSORT Diagrams. Participant screening, enrollment, randomization, infusion completion, follow-up completion, and inclusion in the analyses are shown for the 1 mg/kg cohort **a** and the 3, 10, and 30 mg/kg dose-escalation cohorts **b**. IMP, investigational medicinal product; LLQ, lower limit of quantification; HBsAg, hepatitis B surface antigen; HBeAg, hepatitis B e antigen.

**Supplementary Fig. 2:**
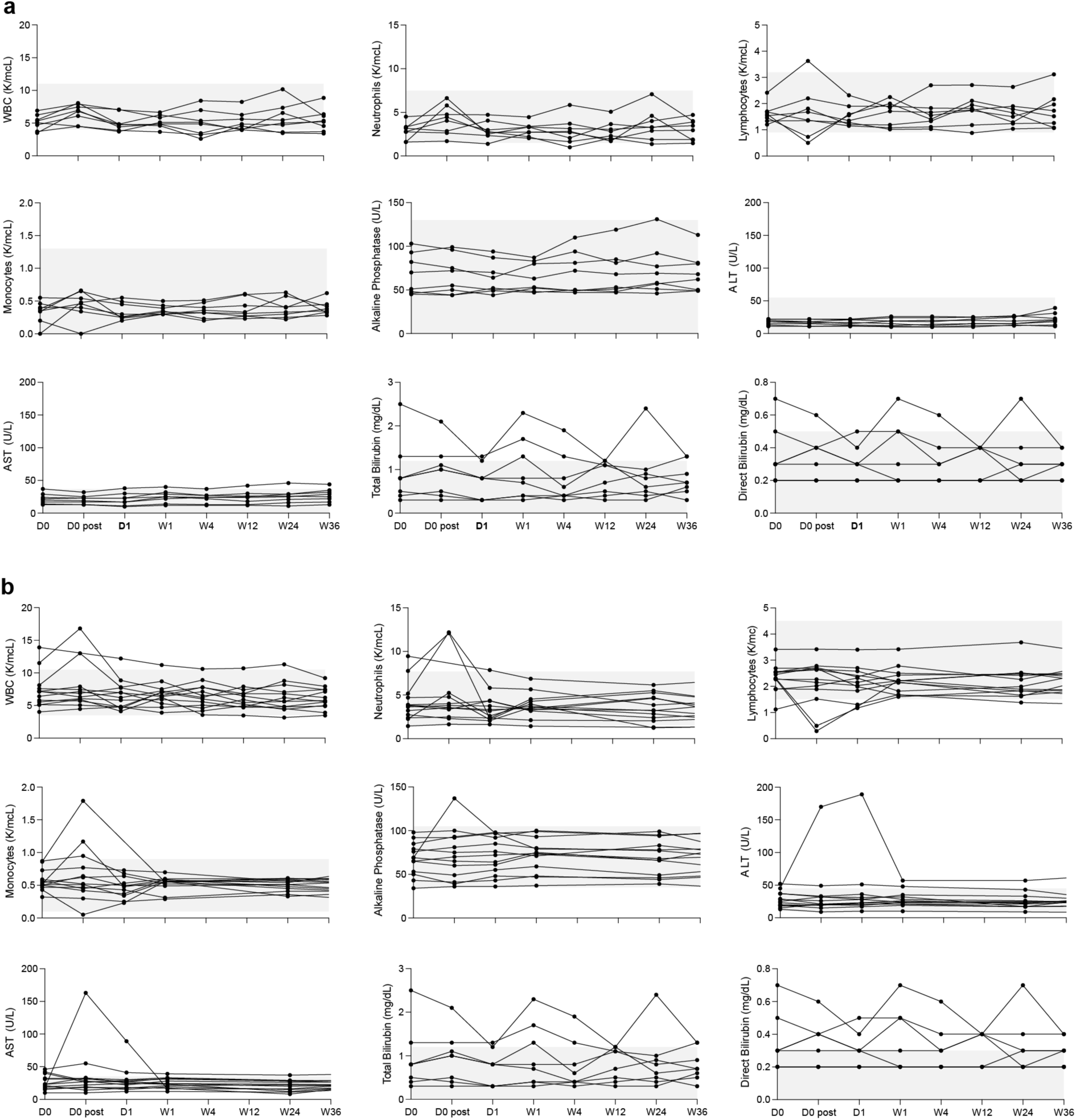
Clinical laboratory results. **a**, US study participants (n=8). **b**, European study participants (n=13). Shaded area illustrates the normal range for the measured parameter.

**Supplementary Fig. 3:**
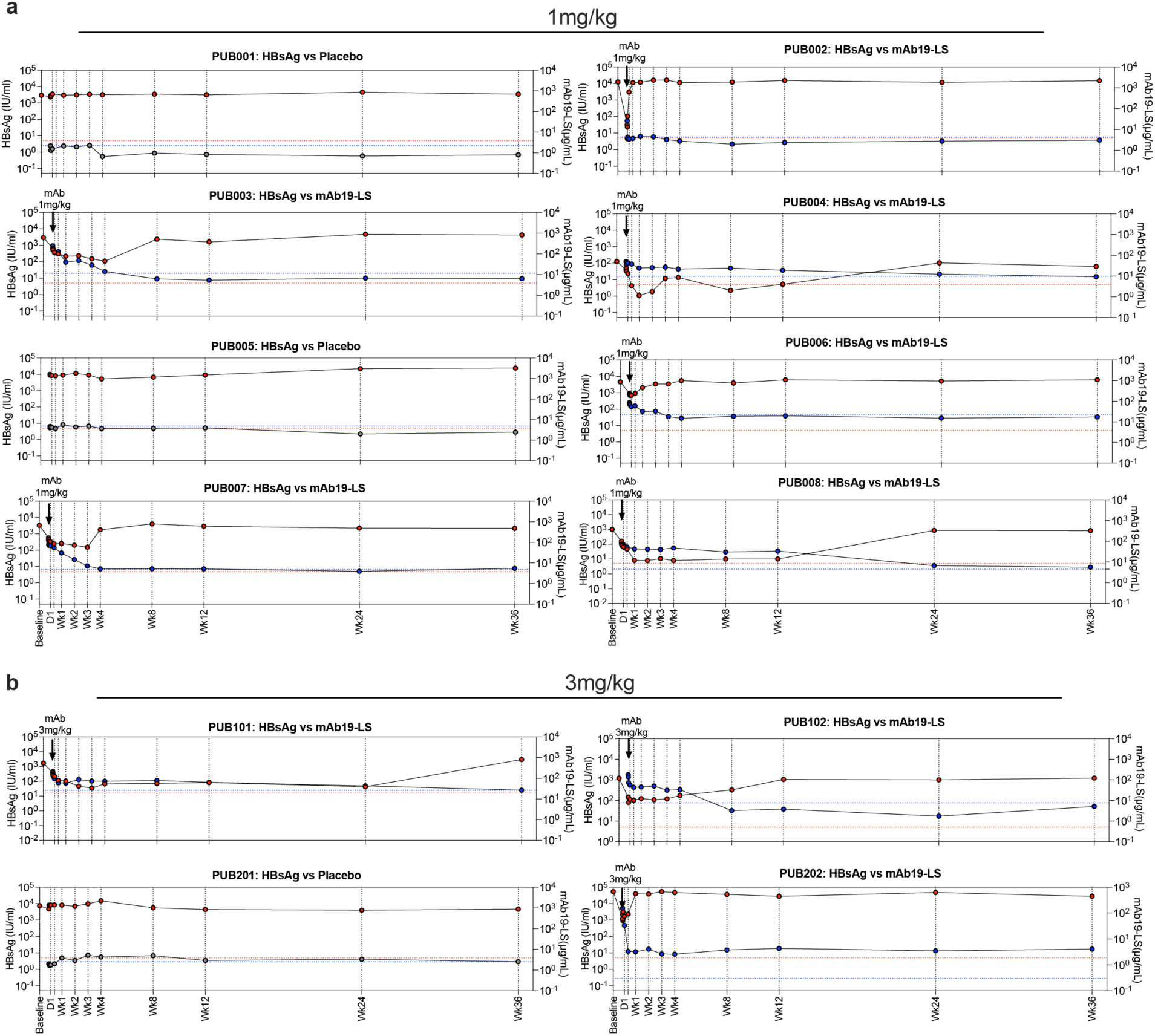
Individual dynamic of mAb19-LS concentration and corresponding HBsAg level. Longitudinal serum mAb19-LS concentrations and HBsAg levels for individual participants over 36 weeks. Antibody concentrations declined over time, with variability associated with baseline antigen burden. Blue indicates mAb19 levels, with dotted blue lines showing background mAb19 at baseline. Red indicates HBsAg levels, with dotted red lines showing LOD for HBsAg quantification.

**Supplementary Fig. 4:**
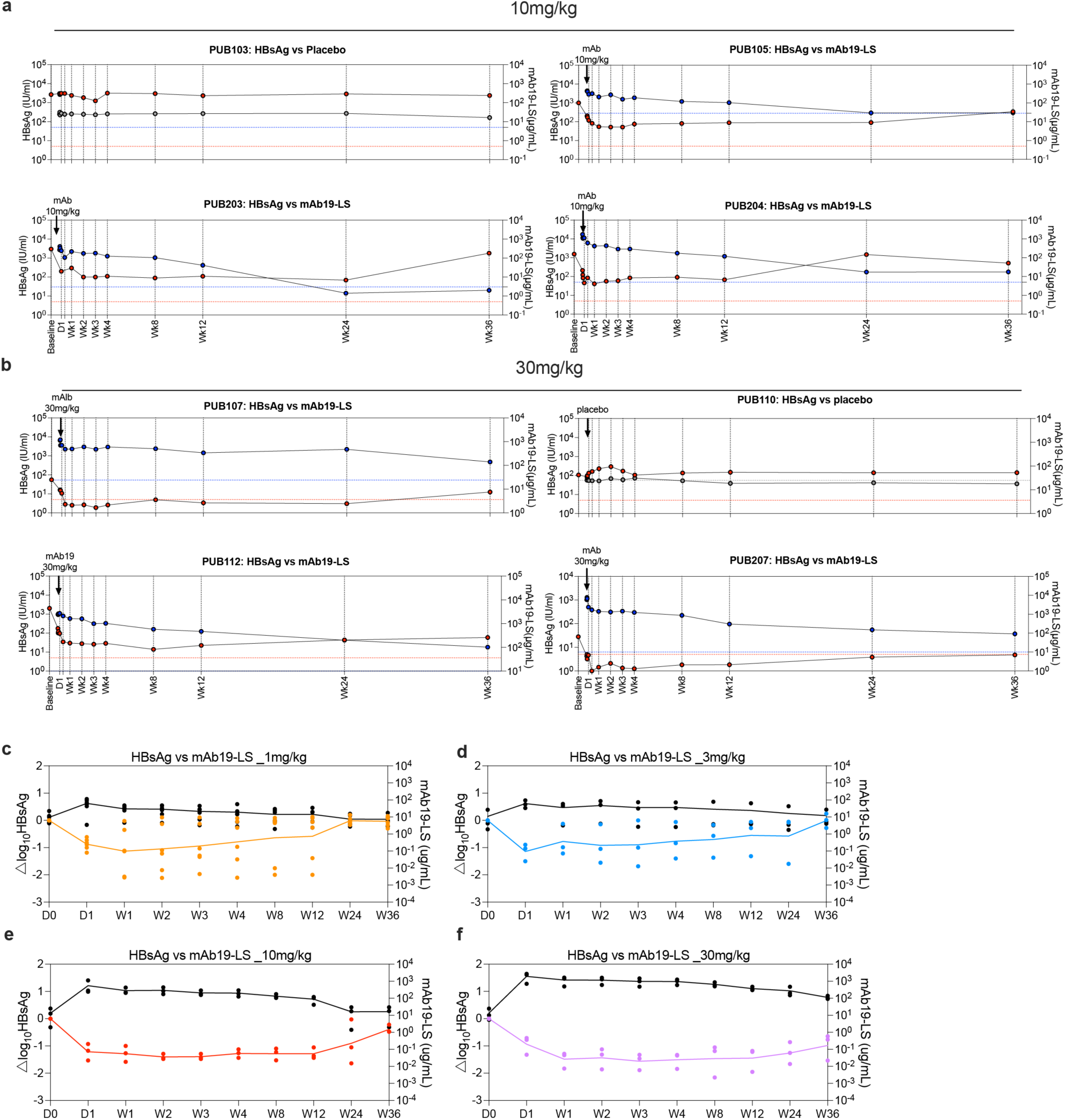
Exposure–response relationship between mAb19-LS concentration and HBsAg reduction. **a,b**, Representative individual plots showing concurrent mAb19-LS concentration and HBsAg levels over time. **c-f**, Mean HBsAg reduction and mean serum mAb19-LS concentration for each dose cohort. Higher antibody exposure was associated with extended antigen suppression. Blue indicates mAb19 levels, with dotted blue lines showing background mAb19 at baseline. Red indicates HBsAg levels, with dotted red lines showing LOD for HBsAg quantification.

**Supplementary Fig. 5:**
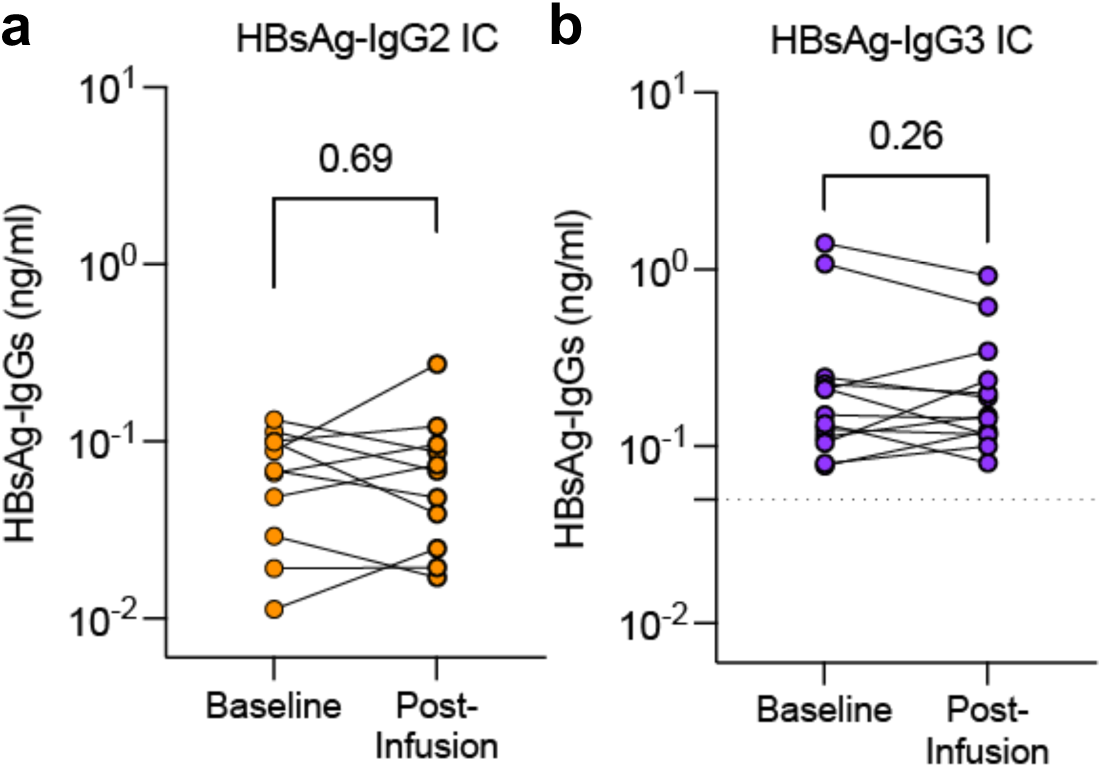
Quantitation of IgG2 and Ig3 within circulating HBsAg–IgG immune complexes. **a,b**, Quantification of HBsAg–IgG2 or HBsAg–IgG3 immune complexes at baseline and post-infusion following PEG 6000 precipitation of circulating immune complexes. Each line represents one participant. Values are shown as concentration (ng/mL). P-values were calculated using paired t-test, n=14.

**Supplementary Fig. 6:**
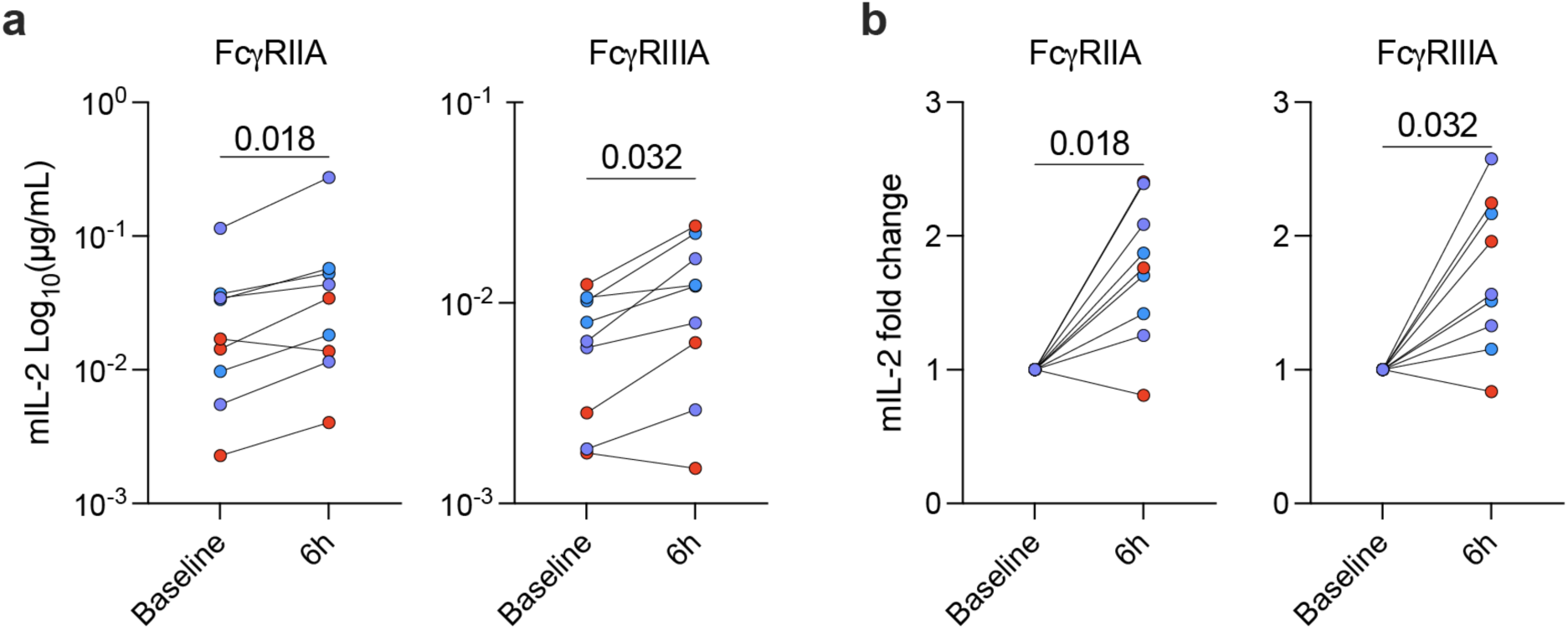
Serum obtained after mAb19-LS infusion activates FcγRIIa and FcγRIIIa reporter cells. **a,b**, Serum samples obtained before infusion and at 6 hours post-infusion from participants receiving 3, 10, or 30 mg/kg mAb19-LS were incubated with IL-2 reporter cells expressing human FcγRIIa or FcγRIIIa. Reporter activation was measured by mIL-2 concentration **a** and fold change **b** relative to baseline. Each point represents one participant; lines connect paired samples. Colored symbols denote individual participants, participants that received 3mg/kg (blue), 10mg/kg (red) and 30mg/kg (purple).

**Supplementary Fig. 7:**
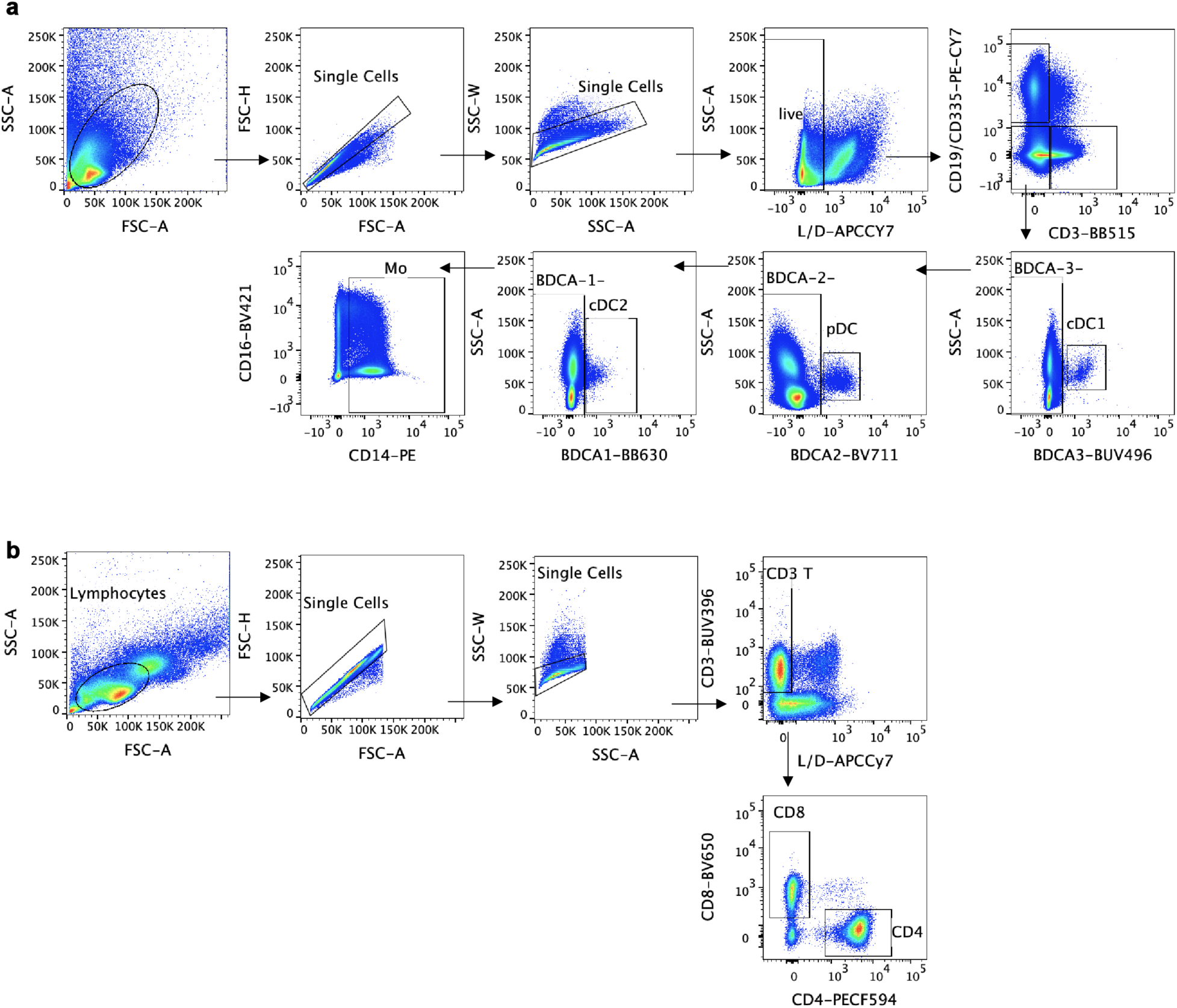
Flow cytometry gating strategy. **a**, Representative gating strategy for identification of circulating DC subsets and monocytes. Cells were first gated on singlets and live cells, followed by exclusion of lineage-positive populations. cDC1, cDC2, pDC, and monocytes were defined based on canonical surface marker expression as indicated. **b**, Gating strategy for T cell panel. Following stimulation, cells were gated on singlets, live cells, CD3⁺ T cells, and then CD4⁺ or CD8⁺ subsets prior to analysis of dextramer staining.

**Supplementary Fig. 8:**
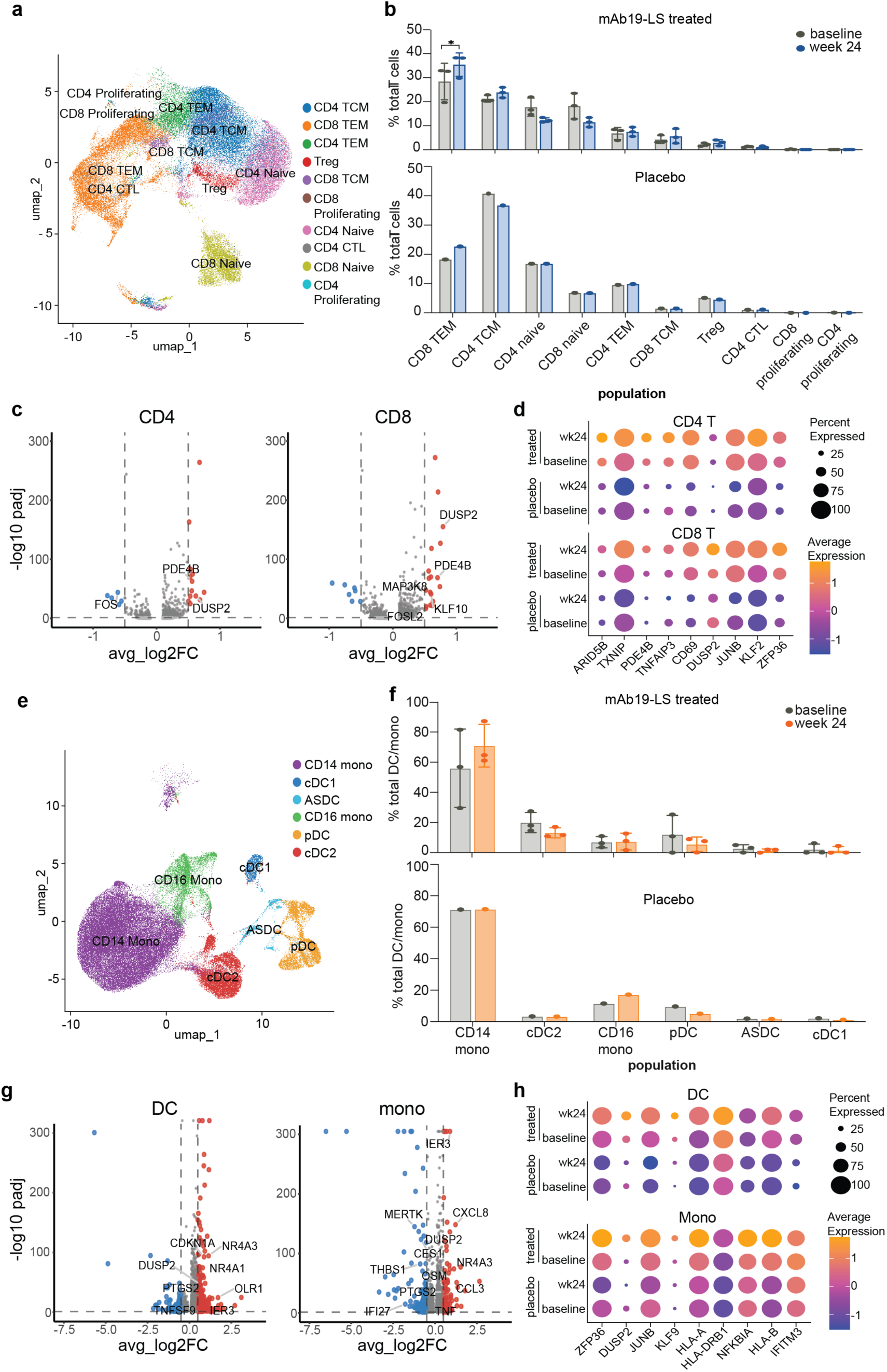
Differentially expressed genes in T cells and APCs in treated participants. **a**, UMAP depicting cells from scRNA-seq on CD4^+^ and CD8^+^ T cells sorted from 3 treated and 1 placebo participant using samples collected pre-infusion (baseline) and at week 24 (Wk24). **b**, Frequency of each T cell subset from A in treated (top) and placebo (bottom) at baseline and week 24. CD8 TEM p-value=0.05. **c**, Volcano plots showing differentially expressed genes between Wk24 and baseline in treated participants, with selected genes highlighted. Red dots indicate genes up-regulated at Wk24, blue dots represent genes down-regulated in Wk24, and grey dots represent non-changed genes. **d**, Expression of the leading-edge features associated with IFN-γ and TNF-α signaling in CD4^+^ (top) and CD8^+^ (bottom) cells from treated and placebo participants at Wk24 compared to baseline. **e**, UMAP of cells from scRNA-seq on DCs and monocytes (mono) sorted from the same participants and timepoints as in **a**. **f**, Frequency of each cell subset from **e** in treated (top) and placebo (bottom) at baseline and Wk24. **g**, Volcano plots showing differentially expressed genes between Wk24 and baseline in treated participants, with selected genes highlighted. Red dots indicate genes up-regulated at Wk24, blue dots represent genes down-regulated in Wk24, and grey dots represent non-changed genes. **h**, Expression of the leading-edge features associated with IFN-γ and TNF-α signaling in DCs (top) and monocytes (bottom) from treated and placebo participants at week 24 compared to baseline. Statistics in **b** indicate paired t-test.

---

Supplementary Table 1. Participant Demographics.

Supplementary Table 2. Safety Summary

Supplementary Table 3. Listing of Reported Adverse events.

Supplementary Table 4. Clinical Laboratory Results (a) Safety labs, (b) Serum HBsAg levels, (c) Other HBV markers.

